# Adverse events and their association with comorbidities after first and second doses of Covishield vaccination among healthcare workers of Government owned medical colleges in Kerala

**DOI:** 10.1101/2021.05.19.21257317

**Authors:** A Remlabeevi, Thomas Mathew, G S Harikumaran Nair, Greeshma Lathika Rajasekharan Nair, Mariyam Rajee Alex

## Abstract

**Background:** A bridging study in the population was not existing at the time of introduction of Covishield vaccine in the state of Kerala A monitoring network for adverse events which was already in place ensured the reporting of serious adverse events following vaccination, but the recording of symptom profile and timeline of symptoms along with the comorbidity status of the individual recipients needed a further database.

**Aims:** To find the proportion of vaccine recipients with adverse events following the first and second doses of Covishield vaccination along with assessment of the symptom profile and timeline of appearance of symptoms following vaccination with each dose along with association of adverse events with comorbidity status of the respondents.

**Materials & Methods:** Cross-sectional study with secondary data taken from the AEFI database of the Covid Cell, Directorate of Medical Education of the Kerala state.The database is formed with responses collected as online self-reporting forms collected from the health workers (doctors, nurses, students, paramedical, housekeeping and clerical staff) who received vaccination from vaccination centres in government owned Medical Colleges in Kerala for a period of three months from the date of rolling out vaccination in the state.

**Results:** A total of 4402 healthworkers submitted the forms after taking the vaccination,either first dose or second dose.Out of this 3656(83.1%)responders were after first dose and 746(16.9%)participants responded after second dose 63.3% respondents after first dose & 24.3% after second dose reported they had experienced adverse events following vaccination with first or second dose of the vaccine respectively.The first symptom to be noticed in those who reported the adverse event after first dose was body ache (17.9%) followed by headache in 15.1 % of participants. 11% (403 out of 3656)of the responders after first dose were having comorbidities and 8.3 % were taking concomitant medications. History of being an asthmatic was found to be of increased risk for developing symptoms following first dose of vaccination(p value 0.004, OR-1.269,95% CI 1.127-1.429) whereas diabetes mellitus is not identified as a risk factor for development of adverse events though a significant association is found,might be due to a decreased reactogenicity.Among those who responded after receiving second dose of vaccination,24.3% reported they had adverse events(at least one post vaccination symptom),of which the first symptom experienced was headache (25.5%),followed by fever(20.9%) as compared to bodyache and headache after the first dose.

**Conclusions:** 56.7% of those who responded after receiving either first or second dose of the vaccine developed at least one symptom afterwards (63.3% after first and 24.3% after second dose of the vaccine respectively)with mean duration of appearance of symptoms being 8.5 hours and for majority of respondents the symptoms lasted for a day only.The first symptom to appear was bodyache (first dose),fever(in second dose) Though 8.5% respondents had a history of previous Covid infection it had no association with adverse events.Symptoms like chestpain,dry mouth, breathing difficulty which are not being spelled out in Covishield factsheet, has also been reported by the study respondents.Seizures were also reported as an adverse event by the responders

---

The emergence of SARS-CoV-2 virus as a public health threat in the year 2020 brought forth a number of varied challenges along with evolution of different platforms and methodologies in the public health crisis management.In December 2019,it was noted that the adults from the capital of Hubei province in Wuhan China began diagnosed with a severe pneumonia of unknown cause Many of the initial cases had a common exposure to the Huanan wholesale seafood market that also traded live animals.Out of 41 initial cases reported two thirds were found to have an exposure to the market.^(1)^ After the outbreak of SARS there was a surveillance system established in the country which became instrumental in collection of respiratory samples and detection of the SARS-CoV-2 virus^.(2)^

The first case of SARS-CoV-2 in the state of Kerala was a medical student who returned from Wuhan,China who was also the first confirmed case of the same in India The first case was reported on 29.1.2020 in Thrissur district and she was admitted to the isolation ward of Govt medical college,Thrissur and treated And then the state entered into the vortex that went on to become the Covid 19 pandemic in the state.The state now reports nearly 1272645 cases, of which 1148671 cases have recovered from the illness and 4978 deaths as in third week of April 2021 ^(3)^ State has been able to tackle the crisis posed by the pandemic to a great extent through a strong surveillance network and through targeted continuum of care services offered by an organized network of covid care centres and hospitals spanning through the primary level to the tertiary care.Of late the fight against the virus is augmented by the mass vaccination provided to the healthcare workforce and also to the population.

The world is now in very much need of effective and safe vaccines against SARS-CoV-2 as the pandemic succeeds to subsequent waves in various parts of the globe.The vaccine development was thought of as a tedious time consuming process but in the very recent times,the scientific community has succeeded in developing safe and effective vaccines against SARS-CoV-2 in a very expeditious manner.There are 89 vaccines now in clinical trials involving human beings of which 23 have entered the final stage. Also nearly 77 vaccine candidates are undergoing preclinical evaluation in various animal studies^(4)^ The rampant spread of SARS-CoV-2 pandemic has led to an urgent race in the development of a vaccine(5)Development of a safe and efficacious vaccine is said to be the most effective approach in prevention of the ongoing pandemic(6). The perceived disease risks are often outweighed by the Vaccine safety concerns when it comes to an individual level decision of whether to take the vaccine or not. Hence, information regarding the safety of vaccine among the public forms an important determinant of the success of vaccination drive(7).

Out of many potential vaccine candidates, ChAdOx1 nCoV-19 vaccine by the Oxford was among the forerunners to enter into human trials and to get documented with reliable safety and efficacy profile. The ChAdOx1 nCoV-19 vaccine (AZD1222) was developed at Oxford University and consists of a replication-deficient chimpanzee adenoviral vector ChAdOx1, containing the SARS-CoV-2 structural surface glycoprotein antigen (spike protein; nCoV-19) gene. Interim analysis of safety and efficacy of the same vaccine from randomized controlled trials held in four countries showed that 175 severe adverse events occurred in 168 participants, 84 events in the ChAdOx1 nCoV-19 group and 91 in the control group(8).Out of the two versions of the vaccines developed,the one developed by the Serum Institute of India,the Covishield vaccine was given as part of mass vaccination strategy in the country but there was no prior bridging study done in the population.

With a population of 1.38 billion, in the initial phase of the COVID-19 vaccination programme, India aimed at vaccinating 300 million people by August 2021, including 30 million health workers and frontline workers (e.g., police, soldiers), and 270 million elderly people (i.e., aged over 50 years) and people with co-morbidities. According to the Press Information Bureau, COVID-19 vaccination in India was initiated with two types of vaccines: Covishield (by Serum Institute of India Ltd) and Covaxin (by Bharat Biotech International Ltd)(9)Later Sputnik V vaccine by Dr Reddy’s has also been approved for emergency use in the country.

## Background and rationale

The state of Kerala started its vaccination to healthcare workers on 16th of January 2021 through Government and private stakeholders and a network of adverse event reporting units were set up along with this in each district. This monitoring network ensured the reporting of serious adverse events following vaccination, but the recording of symptom profile and timeline of symptoms along with the comorbidity status of the individual recipients needed a further database. A survey by Rajeev Jayadevan etal among healthcare workers who received the first dose of Covishield/Covaxin/Pfizer vaccine reported that 65.6% of the recipients had at least one post vaccination symptom.(10). Still the significance of presence of comorbidities and concurrent medications needs to be studied further along with a detailed analysis of symptoms following second dose. The ChAdOx1 nCoV-19 vaccine (AZD1222) vaccine trials also show the incidence of neurological complications 14-21 days post second dose of vaccination. Also, a comparison of symptoms following two doses is being carried out in the present study.

## Objectives of the study

The study was done to find the proportion of vaccine recipients with adverse events following the first and second doses of Covishield vaccination along with assessment of the symptom profile and timeline of appearance of symptoms following vaccination with each dose.A comparison of the symptom profile following first and second doses of the vaccine was also done along with finding out an association between coexistence of comorbidities with presence of adverse events.

## Materials and methods

Cross-sectional study with secondary data taken from the AEFI database of the Covid Cell, Directorate of Medical Education of the Kerala state.The database is formed with responses collected as online self-reporting forms collected from the health workers (doctors, nurses, students, paramedical, housekeeping and clerical staff) who received vaccination from vaccination centres in government owned Medical Colleges in Kerala from January to March 2021. Study variables includes sociodemographic variables, presence of post vaccination symptoms, time of onset and duration of symptoms, comorbidities present, towards vaccine etc.

The study tool used is a self-reporting google form in English and Malayalam, sent from the Covid Cell, Directorate of medical education to the Heads of the institution of individual Medical Colleges. All those who responded at least 72 hours after receiving the vaccine (first or second dose) are taken into the study.Data was entered into excel 2019 and analysed using SPSS v25.

## Results

A total of 4402 healthworkers submitted the forms after taking the vaccination,either first dose or second dose.Out of this 3656(83.1%)responders were after first dose and 746(16.9%)participants responded after second dose The mean age of the study participants is 30 years(SD-11.3years),the youngest was of 18 years and eldest participant of 68 years.71.6% of the study participants were females,the higher participation of females may be due to the nursing population and also increasing number of females in the doctor/medical student population. 50.6% (2229)of the participants are doctors and 25.2% belong to the nursing staff.The other participants belong to dental surgeons, laboratory technicians,pharmacists and other auxiliary staff.There were 3148(71.5%) study participants from 18-35 year agegroup,1214(27.6%)from 36-60 years and 40 persons who responded after vaccination were above 60 years.

Regarding the history of Covid 19 positivity,349 (7.9%) gave a history of previous Covid 19 infection.10.8 % of the study participants belonging to the above 50 year age groups gave a gave a history of having tested positive for Covid 19 previously,but the history of infection was not found to have any significant association with incidence of adverse events in any of the age groups.As a whole 56.7%(2495)respondents reported they had experienced adverse events following vaccination with either first or second dose of the vaccine.

Following the first dose, 63.3% had developed adverse events and they were mainly minor namely,low grade fever,headache,myalgia,fatigue,dizziness etc.A small fraction of the study participants also reported severe symptoms,which is discussed in detail. 8.5% of the respondents (309 out of 3656) reported that they were tested positive at an early point of time before receiving the vaccination.Among them 188 persons have reported that they had experienced one or the other symptom following vaccination wheras 121 persons had no adverse event following vaccination. The first symptom to be noticed in those who reported the adverse event after first dose was body ache (17.9%) followed by headache in 15.1 % of participants.The mean duration of appearance of symptoms is 8.6 hours though it varied from immediately following vaccination to upto 96 hours post vaccination with first dose.Mean duration of having symptoms was 1.7days(SD+-0.9day)Around 46.5 % responded that the symptoms lasted only less than a day, wheraeas 3.6% responded they had one or the other symptom for more than 3days. 11% (403 out of 3656)of the responders after first dose were having comorbidities and 8.3 % were taking concomitant medications.Majority of them reported hypothyroidism as a comorbidity.28 persons reported they had a history of cardiac disease,83 with diabetes mellitus and 68 with bronchial asthma.Also there were 13 persons with history of autoimmune diseases,3 persons with known history of seizure disorder and one with type 1 diabetes mellitus and 4 with a history of carcinoma,all of them with no serious adverse event following vaccination.

**Fig 1:**
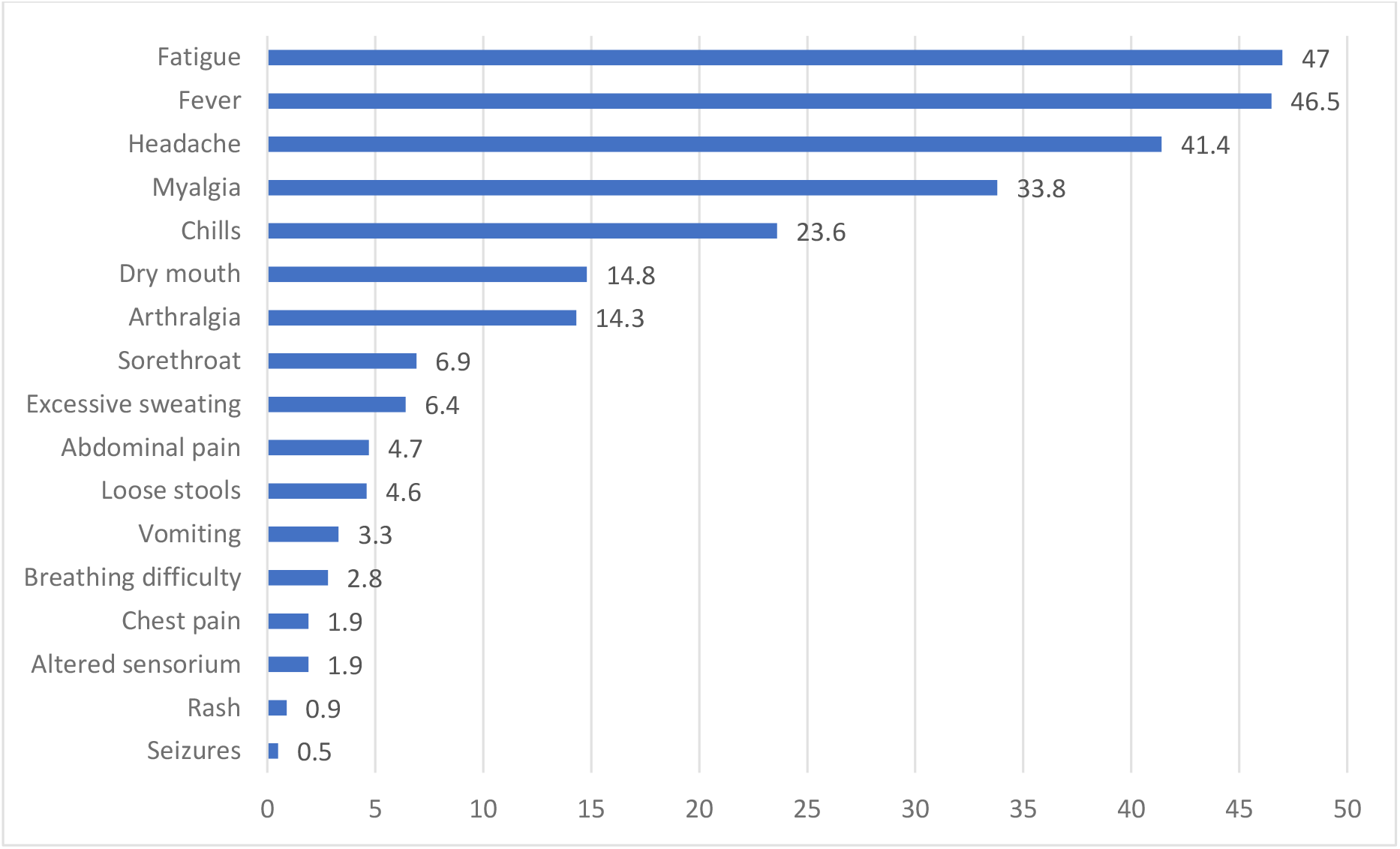
Adverse events following first dose of vaccination (%)

From the results it has been shown that elderly (age more than 60 years) have no significant association with the development of adverse events following the first dose.Older persons are not at increased risk for developing symptoms following vaccination (p value 0.139.OR 0.773,95%CI(0.518-1.154)

**Table 1 :** Age and adverse events following first dose of vaccination

|  | 18-40 years | 41-60years | >60years | P value |
| --- | --- | --- | --- | --- |
| Time of onset of symptoms (hours), mean(SD) | 8.45(±5.096) | 10.05(±8.24) | 12.83(±8.62) | <0.001(sig) |
| Duration of symptoms(days) mean(SD) | 1.68(±.68) | 1.84(±1.05) | 1.5(±0.52) | 0.008(sig) |
| Presence of symptoms following vaccination, n(%) | 1945(67.2) | 354(48.4) | 15(53.5) | <0.000(sig) |

Difference in age groups was significantly associated with the incidence of adverse events following first dose;Higher incidence is noted in 18-40 year age group(this age group has a higher participation in the study population)Also time of onset of symptoms is found to be delayed in elderly.Among all the female participants, 59.9% were reported to have adverse events which is a higher incidence as compared to that among men(48.6%).Females were found to have a higher incidence as well as a significant association with incidence of adverse events after first dose(p value<0.000,OR-1.65,95 %CI 1.429-1.917) and second dose(p value 0.021,OR-1.5, 95% CI 1.028-2.247).

Preexistence of any comorbid illness(Diabetes mellitus,Cardiac disease,Immunological disorder,Hypertension or Bronchial asthma) was not found to be significantly associated with incidence of adverse events either after first or second dose(p value-0.589,OR-1.059,95% CI-0.873-1.283). History of being an asthmatic was found to be of increased risk for developing symptoms following first dose of vaccination(p value 0.004, OR-1.269,95% CI 1.127-1.429) whereas diabetes mellitus is not identified as a risk factor for development of adverse events though a significant association is found.It may be concluded that Diabetes might be causing a less reactogenicity to the vaccine among the recipients as its not identified as a risk inspite of having a significant association with occurrence of adverse events after first dose(p value-0.001,OR-0.647,95% CI 0.496-0.843).Cardiac disease was not found to be significantly associated with incidence of adverse events following first dose. As obtained from multivariate analysis,bronchial asthma is a predictor variable for development of adverse events following first dose of Covishield vaccination (pvalue<.001,95%CI-1.342-4.769).

Among those who responded after receiving second dose of vaccination,24.3% reported they had adverse events(at least one post vaccination symptom),of which the first symptom experienced was headache (25.5%),followed by fever(20.9%) as compared to bodyache and headache after the first dose.The mean duration of onset of symptoms after receiving the second dose was about 8.7 hours with symptoms lasting for a maximum of 120 hours, which is longer as compared to the first dose.Symptoms lasted for a mean duration of 1.8 days,SD+-1.4 days(higher than that of first dose) and maximum duration of symptoms as reported by a participant was 10 days as compared to 2weeks reported earlier after first dose.After the second dose 57.5% of those with adverse events responded that the symptoms lasted for less than a day as compared to 46.5% of first dose respondents.Only 5% participants responded that their symptoms lasted for more than 3days.There were only 5.4% of participants who reported a history of Covid 19 infection and around onethird among them reported the incidence of adverse events which is comparable with that of first dose.Among female participants 26.4% were having adverse events and among males only 19.1%,very low incidence as compared with the first dose which is well expected.Female gender was found to be significantly associated with adverse events following second dose also(p value 0.021,OR-1.5,95% CI-1.028-2.247).But unlike the first dose, the difference in age groups was not found to have any significant association with adverse events.Among those who had adverse events malaise,fatigue,headache,myalgia were the major symptoms, but chest pain was of higher incidence (5%)as compared to the first dose(2.9% in those with adverse events).The incidence of dry mouth decreased (8.8%) as compared with first(23.4%) and 20 persons out of 3656 respondents of first dose had reported seizures as an adverse event whereas after second dose only one person reports to have developed seizures.

Among those who responded after second dose, there were only 70 respondents who had a history of any comorbid illness,out of which only 20 developed adverse events,13 respondents who were asthmatic (5 among them developed adverse events),20 respondents with diabetes mellitus (2 had adverse events)with no significant association between these variables.

## Discussion

The vast majority of adverse events reported after Covid 19 vaccination may be attributed to development of immune response rather than an allergic reaction,but Covid 19 vaccines being new candidates in the vaccine scenario raises an element of risk as well(11) The different ethnic races,gender,agegroups,comorbidity status -all stratifications in the population may have different responses to the newly developed vaccines.An established AEFI monitoring committee may be accountable only to the serious and severe adverse events, but each symptom following vaccination needs to be documented with a well established surveillance system integrated with pharmacovigilance.(11) From this study,the proportion of respondents having reported at least one post vaccination symptoms is 63.3% and 24.3% after first and second doses of Covishield vaccination respectively.In a previous survey by R Jayadevan etal among healthcare workers in India after first dose of vaccination,the incidence of adverse events was reported to be 65.9%, but it was not exclusive to Covishield vaccination,included other vaccine candidates like PfiZer also.But similar to the finding of the former survey,the symptoms were more likely to be reported by females and the incidence of adverse events decreased with age.Its evident from the present study that the incidence of post vaccination symptoms is higher among women as compared to men.Serious adverse events like seizures were also being reported by the participants of the present study.Vaccine efficacy cannot be commented with this type of a study as its difficult to distinguish between symptoms developed due to mounting of an immune response and allergic reaction.Also this study doesn’t involve antibody estimation, hence a survey regarding the symptoms following the first and second doses along with the association of these symptoms with the comorbidity status of the participants is made out.The vaccine efficacy and safety profile of Astrazeneca vaccine as described in the interim analysis of vaccine trials in four countries suggest that 61% efficacy against SARSCoV2 in the standard two dose group with 84 severe adverse events noted in the Vaccine recived group in a 1.3-4.8 month time span.(8)Since it is an online survey,those with adverse events are more likely to respond and hence,the incidence of adverse events may be higher. Recall bias also tends to play a part in this study especially regarding the timeline of appearance of symptoms.

## Ethical considerations

Submission of the google form was voluntary and there was no compulsion regarding responding to the form.Forms were devised in both English and Malayalam for better understanding of all categories of workers.It was stated that the data was taken for surveillance and study purpose only and those with serious/severe adverse events were asked to report to the casuality/outpatient department of the concerned institution. Responses to the google form denotes implied consent of the vaccine recipient and strict confidentiality of the data is maintained.Permission was obtained from the Institutional Ethics Committee of Govt Medical College,Thiruvananthapuram.

## Conclusion

56.7% of those who responded after receiving either first or second dose of the vaccine developed at least one symptom afterwards (63.3% after first and 24.3% after second dose of the vaccine respectively)with mean duration of appearance of symptoms being 8.5 hours and for majority of respondents the symptoms lasted for a day only.The first symptom to appear was bodyache (first dose),fever(in second dose) Though 8.5% respondents had a history of previous Covid infection it had no association with adverse events.Symptoms like chestpain,dry mouth, breathing difficulty which are not being spelled out in Covishield factsheet, has also been reported by the study respondents.Seizures are also reported as an adverse event by the responders.

## Data Availability

Derived data supporting the findings of this study are available from the corresponding author on request.

